# Dysregulated immunity in SARS-CoV-2 infected pregnant women

**DOI:** 10.1101/2020.11.13.20231373

**Authors:** Morgan L. Sherer, Jun Lei, Patrick Creisher, Minyoung Jang, Ramya Reddy, Kristin Voegtline, Sarah Olson, Kirsten Littlefield, Han-Sol Park, Rebecca L. Ursin, Abhinaya Ganesan, Theresa Boyer, Diane M. Brown, Samantha N. Walch, Annukka A. R. Antar, Yukari C. Manabe, Kimberly Jones-Beatty, William Christopher Golden, Andrew J. Satin, Jeanne S. Sheffield, Andrew Pekosz, Sabra L. Klein, Irina Burd

## Abstract

**Importance:** The effects of SARS-CoV-2 infection on immune responses during pregnancy have not been systematically evaluated.

**Objective:** To assess the impact of SARS-CoV-2 infection during pregnancy on inflammatory and humoral responses in maternal and fetal samples and compare antibody responses to SARS-CoV-2 among pregnant and non-pregnant women.

**Design:** Immune responses to SARS-CoV-2 were analyzed using samples from pregnant and non-pregnant women who had either tested positive or negative for SARS-CoV-2. We measured, proinflammatory and placental cytokine mRNAs, neonatal Fc receptor (FcRn) receptor expression, and tetanus antibody transfer in maternal and cord blood samples. Additionally, we measured anti-spike (S) IgG, anti-S-receptor binding domain (RBD) IgG, and neutralizing antibody (nAb) responses to SARS-CoV-2 in serum or plasma collected from non-pregnant women, pregnant women, and cord blood.

**Setting:** Johns Hopkins Hospital (JHH)

**Participants:** Pregnant women were recruited through JHH outpatient obstetric clinics and the JHH Labor & Delivery unit. Non-pregnant women were recruited after receiving outpatient SARS-CoV-2 testing within Johns Hopkins Health System, USA. Adult non-pregnant women with positive RT-PCR results for SARS-CoV-2, within the age range of 18-48 years, were included in the study.

**Exposures:** SARS-CoV-2

**Main Outcomes and Measures:** Participant demographic characteristics, antibody titers, cytokine mRNA expression, and FcRn receptor expression.

**Results:** SARS-COV-2 positive pregnant women expressed more *IL1β*, but not *IL6*, in blood samples collected within 14 days versus > 14 days after a confirmed SARS-CoV-2 test, with similar patterns observed in the fetal side of placentas, particularly among asymptomatic pregnant women. Pregnant women with confirmed SARS-CoV-2 infection also had reduced anti-S-RBD IgG titers and were less likely to have detectable nAb as compared with non-pregnant women. Although SARS-CoV-2 infection did not disrupt FcRn expression in the placenta, maternal transfer of nAb was inhibited by SARS-CoV-2 infection during pregnancy.

**Conclusions and Relevance:** SARS-CoV-2 infection during pregnancy was characterized by placental inflammation and reduced antiviral antibody responses, which may impact the efficacy of COVID-19 therapeutics in pregnancy. The long-term implications of placental inflammation for neonatal health also requires greater consideration.

---

The ongoing coronavirus disease 2019 (COVID-19) pandemic, caused by severe acute respiratory syndrome coronavirus 2 (SARS-CoV-2), has resulted in over 40 million infections and over one million deaths worldwide^1^. Despite global efforts to characterize the pathogenesis of SARS-CoV-2 infection, the effects of infection on immunity during pregnancy remain undefined. Due to pregnancy-associated immune and endocrine fluctuations, pregnant women and their fetuses are at greater risk for severe complications caused by infectious diseases^2^. Current observations suggest that most pregnant women with COVID-19 are asymptomatic or experience mild disease. The U.S. Center for Disease Control (CDC), however, reports that one in four women, aged 15–49 years, hospitalized for COVID-19 during March 1–August 22, 2020 were pregnant, and these women were more likely to require mechanical ventilation compared to nonpregnant women^3^. The CDC also reports that women infected with SARS-CoV-2 during pregnancy are at higher risk for preterm birth^4^. Because maternal immune activation can be associated with adverse fetal outcomes, including preterm birth ^5,6^, it is possible that SARS-CoV-2 during pregnancy may have detrimental effects on the developing fetus.

During pregnancy, the immune response to viral infection includes secretion of proinflammatory cytokines, such as IL-1β and IL-6, not only at the site of infection but in the placenta as well; these cytokines can readily enter the amniotic cavity and interfere with normal fetal development^5,6^. Thus, even in the absence of severe maternal symptoms or fetal viral infection, the maternal immune response to SARS-CoV-2 could lead to short and long-term consequences in the fetus and neonate, including multiorgan system damage and a predisposition for adverse developmental outcomes^2,7–9^. At the same time, the maternal immune response can also have a protective effect on neonatal health, including the placental Fc receptor (FcRn)-mediated transfer of SARS-CoV-2-specific antibodies transplacentally^10,11^.

In the present study, we investigated immune responses to SARS-CoV-2 using maternal blood, cord blood, and placenta samples collected from pregnant women who had either tested positive or negative for SARS-CoV-2 prior to admission and delivery at the Johns Hopkins Hospital (JHH). We measured maternal and cord blood serum or plasma anti-spike (S) and anti-S-receptor binding domain (RBD) IgG and neutralizing antibody (nAb) responses to SARS-CoV-2, whole blood proinflammatory cytokine mRNA expression, as well as placental cytokine and FcRn expression. Furthermore, we compared antibody responses to an outpatient non-pregnant cohort of women with confirmed COVID-19.

## Results

### Cohorts

Two cohorts were included in this study: the pregnant cohort, consisting of 33 pregnant women who either tested positive (n=22) or negative (n=11) for SARS-CoV-2 prior to delivery (in inpatient or Labor and Delivery settings) at the JHH; and the non-pregnant cohort, consisting of women within reproductive age (18-48 years of age), as defined by WHO ^12^ (n=17) who tested positive for SARS-CoV-2 at an outpatient clinical testing site within the JHH Health System.

Comparing demographic characteristics between SARS-CoV-2 positive and negative pregnant women revealed differences in maternal age at delivery, race, and ethnicity. SARS-CoV-2 positive pregnant women gave birth at a younger age (median =27; IQR 23-34) compared to SARS-CoV-2 negative pregnant women (median =32; IQR 29-35) (*p*<0.05, **Table 1**), were more likely to identify as Other (63.64%) or Black/African American (22.73%) than SARS-CoV-2 negative pregnant women (*p*<0.001, **Table 1**), and were more likely to identify as being of Hispanic/Latina ethnicity (50%) (*p*<0.05; **Table 1**). No significant differences were found between SARS-CoV-2 positive and negative pregnant participants in pre-pregnancy BMI, BMI at delivery, gestational age at birth, neonate late-onset sepsis, chorioamnionitis, time between membrane rupture and delivery, preeclampsia, gestational diabetes, delivery type (cesarean vs vaginal), size of neonate, sex of neonate, NICU stay, or neonatal readmission (**eTable 1**). In comparing SARS-CoV-2 positive pregnant women and non-pregnant women, pregnant women were younger (pregnant median age =27 IQR 23-34; non-pregnant median age =34 IQR 28-41) (*p*<0.05; **Table 1**), less likely to identify as White/Caucasian (14% vs. 47%) (*p*<0.05; **Table 1**), and more likely to identify as Hispanic or Latina (50% vs. 6%) (*p*<0.05; **Table 1**) than non-pregnant women.

**Table 1.** Demographic data SARS-CoV-2 pregnant and non-pregnant cohorts.

|  | Pregnant Female Cohort |  |  |  | Pregnant vs Non-pregnant Female Cohort |  |  |
| --- | --- | --- | --- | --- | --- | --- | --- |
|  | All | SARS-CoV-2 (+) | SARS-CoV-2 (-) | P value * | SARS-CoV-2 (+) Pregnant | SARS-CoV-2 (+) Nonpregnant | P value |
| N (%) | 33 | 22 (66.67) | 11 (33.33) |  | 22 | 17 |  |
| Median Maternal Age at Delivery | 29 | 27 | 32 | 0.0421 | 27 | 34 | 0.006 |
| Race - n (%) |  |  |  |  |  |  |  |
| Asian | 2 (6.06) | 0 (0) | 2 (18.8) | <0.0001 | 0 (0) | 0 (0) | 0.011 |
| Black or African American | 6 (18.18) | 5 (22.73) | 1 (9.09) |  | 5 (22.73) | 6 (35.29) |  |
| Other | 14 (42.42) | 14 (63.64) | 0 (0) |  | 14 (63.64) | 3 (17.65) |  |
| White or Caucasian | 11 (33.33) | 3 (13.64) | 8 (72.73) |  | 3 (13.64) | 8 (47.06) |  |
| Ethnicity - n (%) |  |  |  |  |  |  |  |
| Hispanic or Latina | 12 (36.36) | 11 (50) | 1 (9.09) | 0.0273 | 11 (50) | 1 (5.88) | 0.003 |
| Not Hispanic or Latina | 21 (63.64) | 11 (50) | 10 (90.91) |  | 11 (50) | 16 (94.12) |  |

### Cytokine expression after SARS-CoV-2 infection during pregnancy

Increased inflammation caused by infection during pregnancy can be detrimental for long-term fetal and neonatal outcomes^2,9,13^. We assayed cytokine mRNA expression during SARS-CoV-2 infection as a biomarker for inflammation. Because IL-1β activation during pregnancy can cause adverse fetal outcomes^2,14,15^, we measured *IL1β* mRNA expression in maternal blood, cord blood, and the maternal and fetal sides of placentas, which did not differ between SARS-CoV-2 positive and negative pregnant women (**Figure 1A-D**). To test whether *IL1β* expression differed as a function of maternal symptoms, samples were further categorized based on whether pregnant women were asymptomatic, symptomatic, or SARS-CoV-2 negative. There was a pattern of greater *IL1β* expression in samples from asymptomatic pregnant women (**Figure 1E-G**), with a significant increase in the fetal side of placenta from asymptomatic patients compared to symptomatic patients (*p*<0.05; **Figure 1H**). To assess whether the expression of *IL1β* differed depending on the number of days between a pregnant woman’s PCR test and blood sample collection, maternal blood *IL1β* mRNA expression within each symptom category were compared based on the time window between diagnosis and blood collection. Day 14 was chosen for analysis based on the incubation period of SARS-CoV-2, which extends to 14 days after symptom onset^16^. *IL1β* expression in maternal blood was higher in samples collected within 14 days of a positive SARS-CoV-2 test compared with samples collected > 14 days after test, regardless of symptoms (*p*<0.05; **Figure 1I**).

**Figure 1:**
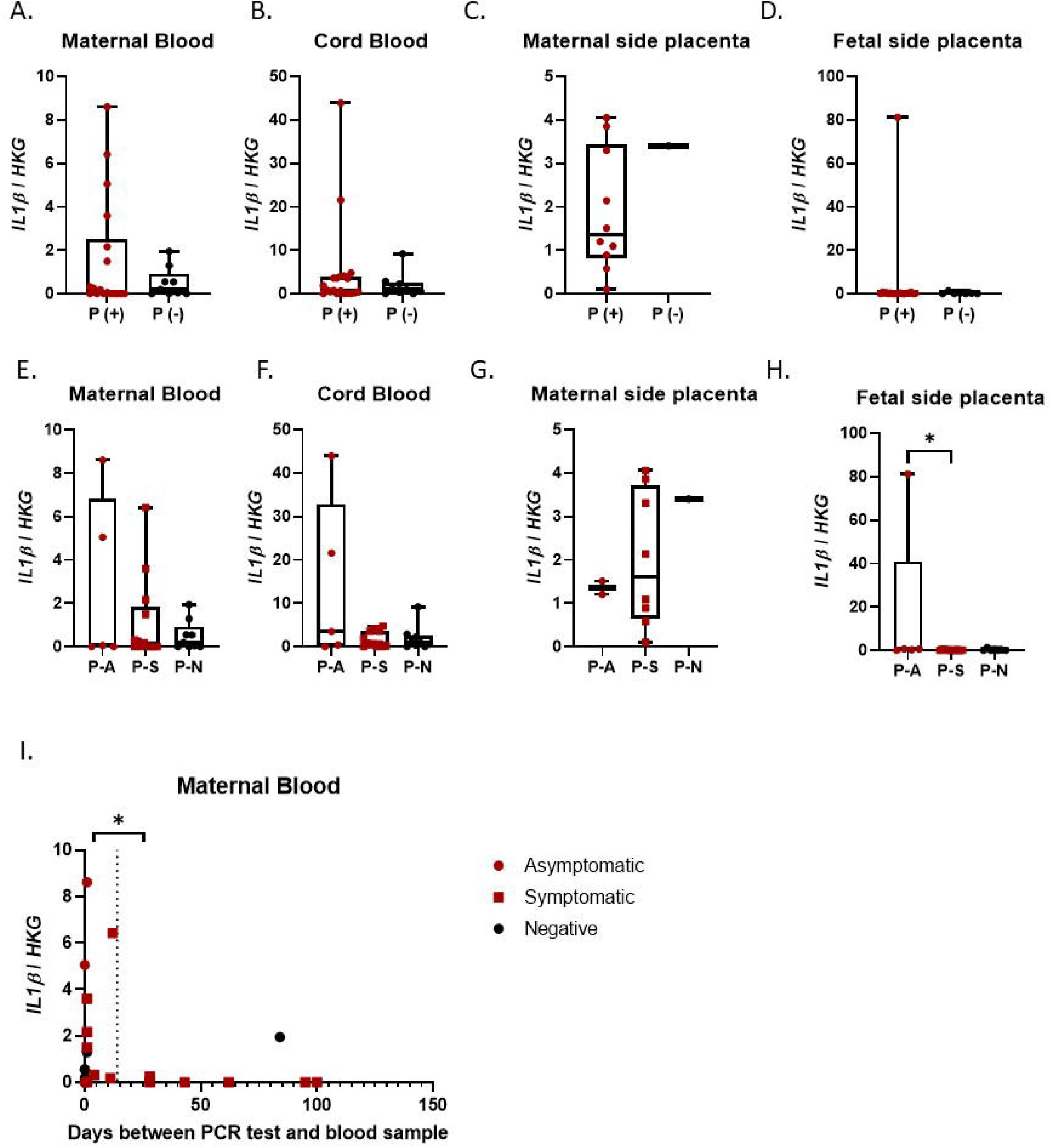
*IL1β* expression in maternal and fetal samples. Maternal and fetal blood and placentas were used to detect *IL1β* gene expression relative to the housekeeping genes (HKG), *18S* and *ACTB*. (**A-D**) Maternal blood, cord blood, and maternal and fetal side placental *IL1β* expression between SARS-CoV-2 positive (P(+)) and negative (P(-)) samples in the pregnant cohort. (**E-H**) Maternal blood, cord blood, and maternal and fetal side placental *IL1β* expression in pregnant women who were asymptomatic (P-A), symptomatic (P-S), or SARS-CoV-2 negative (P-N). (**I**) Maternal blood *IL1β* expression analyzed as a function of symptom expression and days between SARS-CoV-2 PCR positive test and blood sample collection; dashed line located at Day 14; significance denotes comparison of samples collected within 14 days of a positive SARS-CoV-2 test with samples collected > 14 days after test. Maternal blood n= 27; cord blood=29; maternal side placenta n=11; fetal side placenta n=26. \**p*<0.05 by Kruskal-Wallis, Dunn’s multiple comparisons or Mann-Whitney test.

We measured *IL6* mRNA expression in maternal and fetal blood and tissue from our SARS-CoV-2 positive and negative pregnant cohort. It is important to note that all SARS-CoV-2 positive women experienced mild to moderate disease from SARS-CoV-2 infection. In contrast to the elevation observed among severe COVID-19 cases in non-pregnant individuals^17–19^, there was no change in the expression of *IL6* in blood or placentas based on SARS-CoV-2 infection status **(eFigure 1A-D)**, symptom status **(eFigure 1E-H)**, or duration of time between a positive SARS-CoV-2 test and sample collection **(eFigure 1I)**. These data provide evidence of selective *IL1β* mRNA upregulation, particularly early after infection and on the fetal side of the placenta in non-severely ill pregnant women with SARS-CoV-2 infection.

### Antibody responses to SARS-CoV-2 in pregnant and non-pregnant women

To evaluate the impact of pregnancy on immune responses to SARS-CoV-2, antibody responses measured in serum or plasma samples were collected at a median of 34 (IQR: 31.5 – 40) days since confirmed infection, from pregnant (18.91±29.57 days post confirmed infection) and non-pregnant (37.29±12.66 days post confirmed infection) women who had tested positive for SARS-CoV-2. Pregnant and non-pregnant women showed similar titration of IgG (i.e., area under the curve [AUC]) recognizing the full-length SARS-CoV-2 spike (S) protein (**Figure 2A**). In contrast, pregnant women had significantly lower anti-S-RBD IgG titers than non-pregnant women (*p*<0.05, **Figure 2B**). Titers of nAb, however, which correlate with anti-S-RBD antibodies^20^, were measured and were not significantly different between pregnant and non-pregnant women (**Figure 2C**). We also observed that significantly fewer pregnant women (8/17) had detectable nAb titers (i.e., ≥ 1:20 titer) compared with non-pregnant women (16/17) (*p*<0.05; **Figure 2C**), indicating reduced production of neutralizing antibodies in a subset of pregnant women.

**Figure 2.**
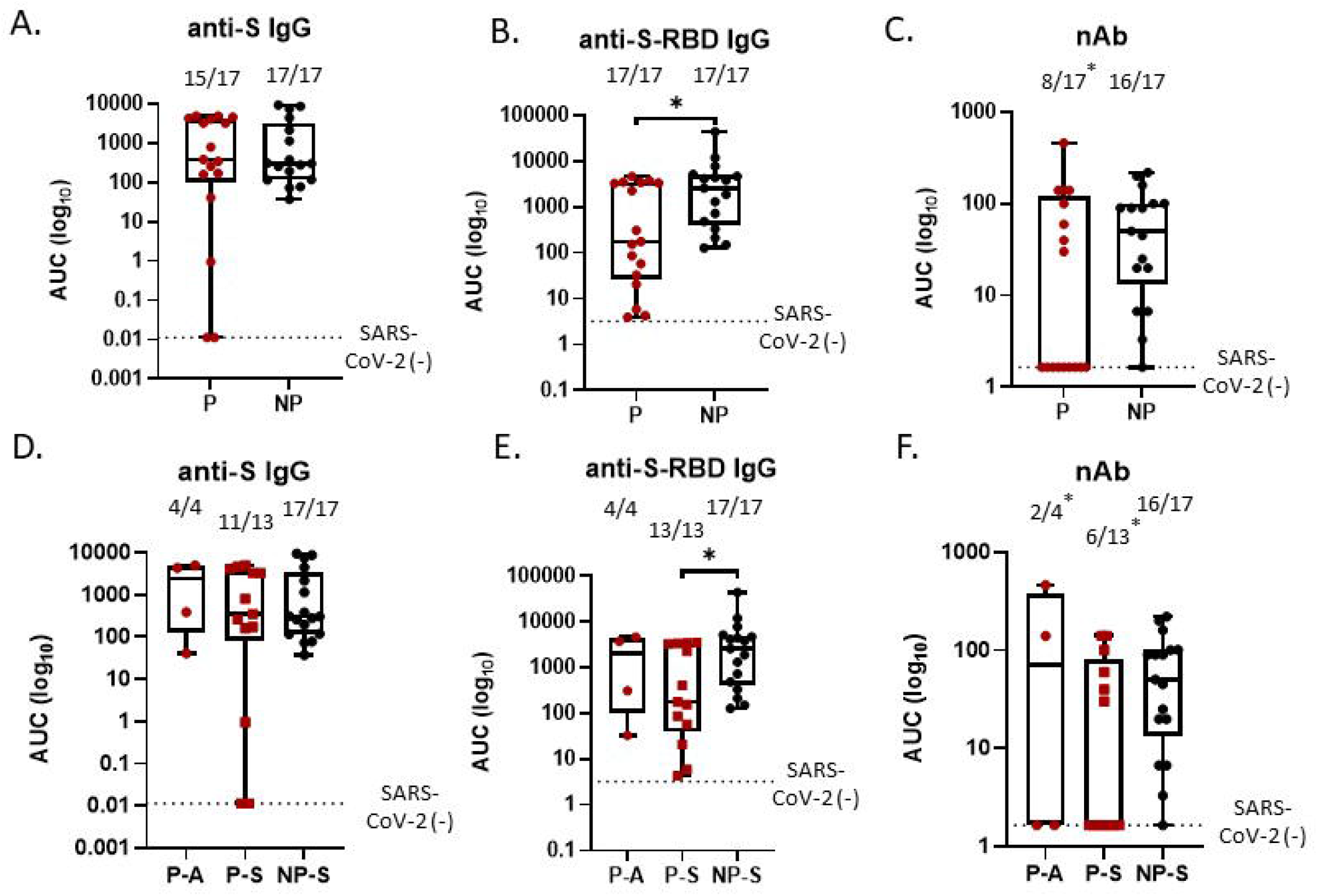
Anti-SARS-CoV-2 antibody titration in samples collected from pregnant and non-pregnant women. Peripheral serum or plasma was used to titer IgG antibodies against SARS-CoV-2 full-length spike (S), S-receptor binding domain (RBD), as well as whole virus neutralizing antibodies (nAb). (A) Anti-S IgG, (B) anti-S-RBD IgG, and (C) nAb area under the curve (AUC) titrations in serum or plasma from pregnant (P) and non-pregnant (NP) women. (D) Anti-S IgG, (E) anti-S-RBD, and (F) nAb AUC titration of peripheral sera from pregnant women characterized as asymptomatic (P-A) or symptomatic (P-S) as well as from non-pregnant symptomatic (NP-S) women. The dashed line denotes the median AUC for SARS-CoV-2 negative samples. Above each box-plot is the proportion of samples with detectable antibody; \**p*<0.05 by Kruskal-Wallis, Dunn’s multiple comparisons, Wilcoxon exact, or Chi-square tests.

To assess whether the presence of COVID-19 symptoms was linked to antibody production, antibody responses were analyzed based on symptom categories. The only antibody response that was influenced by COVID-19 symptoms during pregnancy was maternal anti-S-RBD IgG titers, which were lower in symptomatic, but not asymptomatic, pregnant women as compared with non-pregnant women (*p*<0.05; **Figure 2D-F**). Although overall nAb titers were not influenced by COVID-19 symptoms, both symptomatic (6/13) and asymptomatic (2/4) pregnant women were less likely to have detectable nAb than non-pregnant, symptomatic (16/17) women (*p*<0.05, **Figure 2F**).

To further explore how pregnancy altered the relationship between anti-S-RBD IgG and nAb, titers were directly compared and revealed that anti-S-RBD IgG titers were higher than nAb titers in both pregnant and non-pregnant women (*p*<0.001 **Figure 3A**,**B**). Among pregnant women only, a dichotomy in nAb titers was evident. Consistent with this observation, anti-S-RBD IgG titers in pregnant women with nAb titers <1:20 (i.e., no detectable nAb) were significantly lower than anti-S-RBD IgG titers among pregnant women with nAb titers ≥ 1:20 (i.e., detectable nAb; *p*<0.05; **Figure 3A**). To determine whether time since a SARS-CoV-2 positive test or time since symptom onset could predict antibody responses, we analyzed responses over time. Variation in anti-S-RBD IgG or nAb responses among pregnant women with non-detectable as compared with detectable nAb titers could not be explained by the length of time since a positive SARS-CoV-2 positive test (**Figure 3C**,**D**). Furthermore, time since symptom onset did not explain variation in anti-S-RBD IgG or nAb responses among pregnant women with non-detectable as compared with detectable nAb titers (**Figure 3E**,**F**). Differences in the number of days between a PCR+ test or symptom onset and sample collection also did not statistically explain variation in either anti-S-RBD IgG or nAb responses between pregnant and non-pregnant women (**Figure 3C-F**). These data suggest that, independent of time, pregnancy may reduce the quality of antiviral antibodies against SARS-CoV-2.

**Figure 3.**
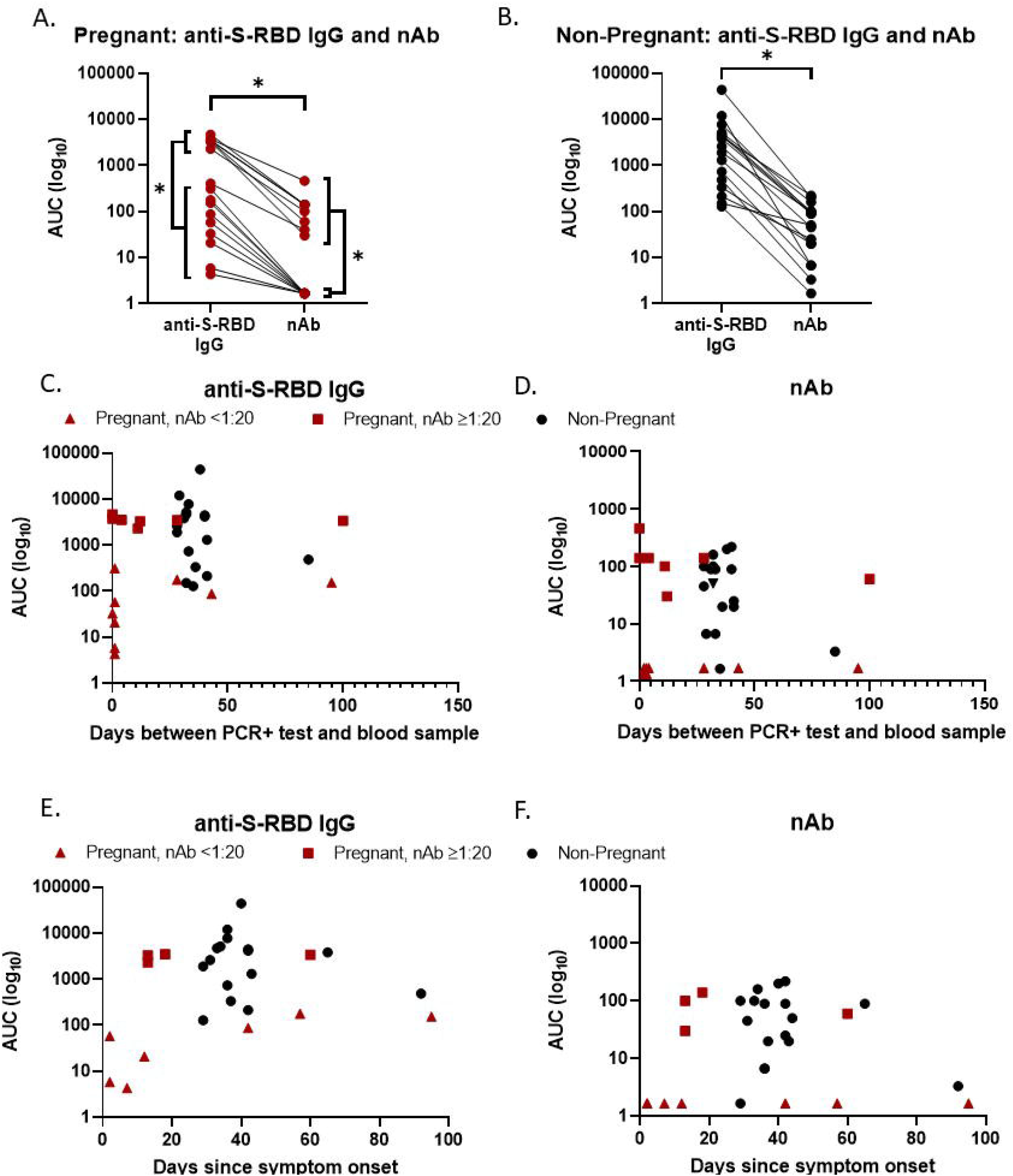
Association between anti-Spike-receptor binding domain (S-RBD) IgG and neutralizing antibody (nAb) titers in pregnant and non-pregnant women. (A) Comparison between anti-S-RBD IgG and nAb AUC in pregnant women, with additional comparison of anti-S-RBD IgG and nAb responses between pregnant with (nAb titer ≥1:20) and without (nAb titers <1:20) detectable nAb. (B) Comparison between anti-S-RBD IgG and nAb AUC in non-pregnant women. (C,D) Anti-S-RBD IgG AUC and nAb analyzed as a function of detectability of nAb and days between SARS-CoV-2 PCR positive test and blood sample. (E,F) Anti-S-RBD IgG AUC and nAb analyzed as a function of detectability of nAb and days since symptom onset and blood collection; missing data points due to unknown symptom onset date n=4. \**p*<0.05 by Wilcoxon exact.

### Antibody transfer in SARS-CoV-2 infection

To assess whether antibody transfer from mother to fetus was affected by SARS-CoV-2 infection, SARS-CoV-2-specific antibody levels in maternal and cord blood serum, FcRn expression, and anti-tetanus IgG titers were assessed in SARS-CoV-2 positive and negative women. Anti-S and anti-S-RBD IgG titers did not differ between maternal and cord blood serum samples (**Figure 4A**,**B**); titers of nAb in maternal serum were, however, significantly greater than in cord blood serum (*p* < 0.05; **Figure 4C**). Protein concentrations of placental FcRn, used as a biomarker of IgG transfer, were not affected by either SARS-CoV-2 infection during pregnancy or symptomatology **(Figure 4D**,**E)**. To further evaluate whether SARS-CoV-2 infection altered the transfer of other antibodies from mother to fetus, maternal and cord blood serum anti-tetanus IgG titers were measured and were not inhibited by SARS-CoV-2 infection during pregnancy **(Figure 4F,G)**. These data suggest that while maternal transfer of nAb may be reduced, SARS-CoV-2 infection does not broadly impact maternal transfer of humoral immunity.

**Figure 4.**
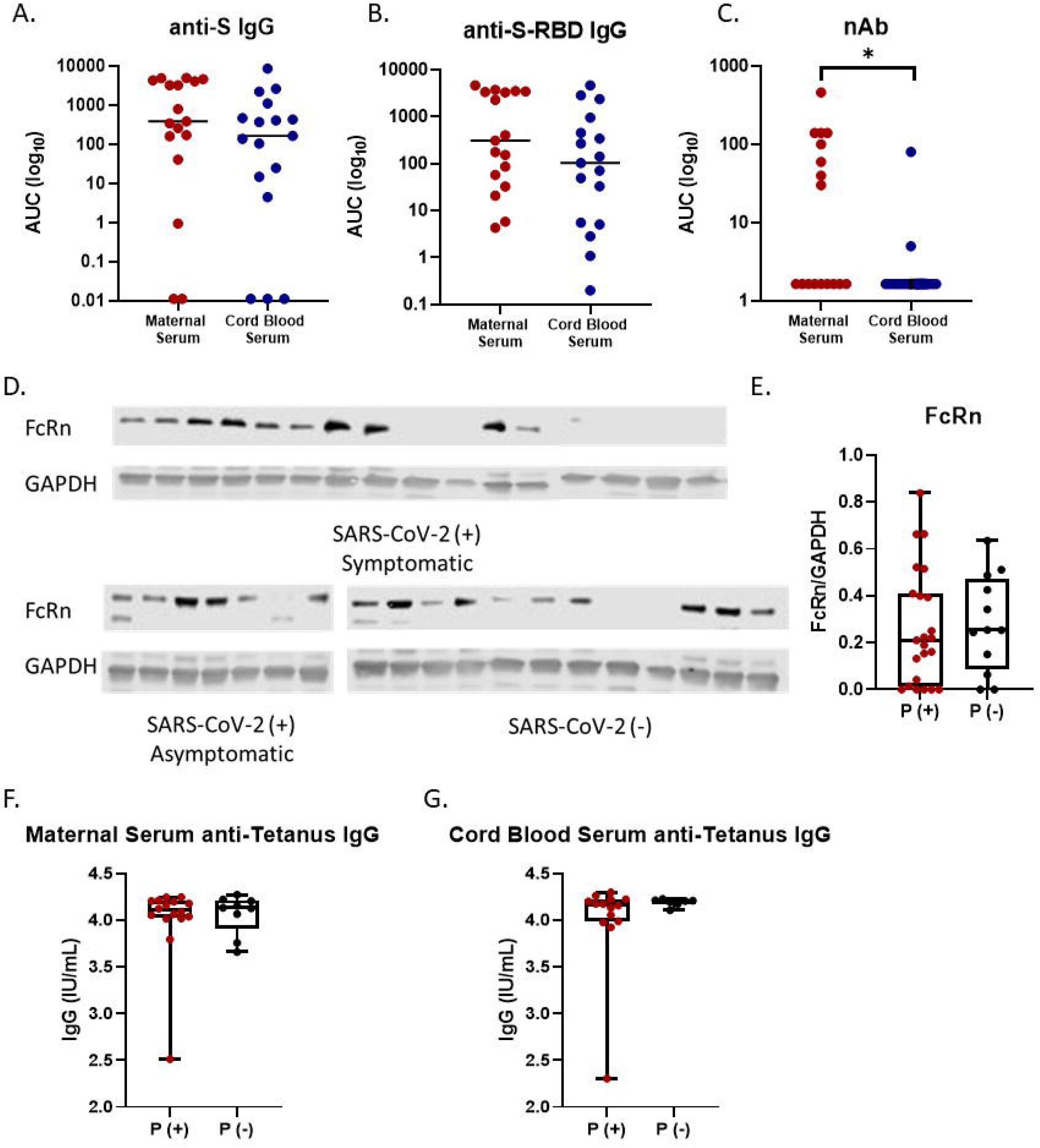
Effects of SARS-CoV-2 infection on antibody transfer from mother to fetus. (A) Anti-S IgG, (B) anti-S-RBD IgG, and (C) nAb area under the curve (AUC) titrations in maternal serum and cord blood serum in SARS-CoV-2 positive pregnant women. (D) Western blot analysis for the neonatal Fc receptor (FcRn) protein in placentas from SARS-CoV-2 (+) symptomatic, SARS-CoV-2 (+) asymptomatic, and SARS-CoV-2 (-) women, n=35. (E) Quantification of FcRn western blot analysis relative to GAPDH was analyzed in placentas from SARS-CoV-2 positive (P(+)) and negative (P(-)) women, n=35. (F) Maternal and (G) cord blood serum anti-tetanus IgG titers in SARS-CoV-2 positive and negative samples in the pregnant cohort; maternal serum n=35, cord blood serum n=21.

## Discussion

Our study provides preliminary evidence that pregnant women exhibit an early inflammatory response and a reduced antiviral antibody response against SARS-CoV-2 as compared with non-pregnant women. The inflammatory response of pregnant women who experienced mild to moderate COVID-19 was characterized by greater *IL-1*β, but not *IL-6*, mRNA expression as has been reported in severe male and non-pregnant female COVID-19 patients^18,19^, as well as greater *IL-1*β mRNA expression on the fetal side of the placenta. Current studies highlight differences in clinical manifestations between SARS-CoV-2 positive pregnant and non-pregnant women, with some studies reporting differences in presenting symptoms, such as lower incidence of fever and cough in pregnant women^21,22^. There is growing evidence that SARS-CoV-2 infected pregnant women face greater risk of hospitalization, intensive care unit admission, and invasive ventilation compared to non-pregnant women, but experience comparable or even lower risk of mortality^23–28^. Studies in SARS-CoV-2 positive pregnant and non-pregnant women report higher frequencies of neutrophils and D-dimer concentrations and lower percentages of lymphocytes, CD4+/CD8+ ratios, and IgG levels in pregnant than non-pregnant women infected with SARS-CoV-2^29–32^. Thus, our study adds to the growing literature demonstrating enhanced inflammatory responses and reduced adaptive immune responses during SARS-CoV-2 infection of pregnant compared to non-pregnant women.

Epidemiological and animal model studies have established the relationship between maternal and placental inflammation and neurodevelopmental disorders, including autism spectrum disorder and schizophrenia, in children^8^. Several proinflammatory cytokines have been identified as mediators of this pathway, responsible for traversing the placenta and triggering neurotoxicity in the developing fetus^33^. Specifically, maternal IL-1 receptor blockade appears to protect against fetal cortical injury in mouse models by attenuating microglial activation, suggesting that IL-1β secretion and signaling play a key role in fetal cortical injury and potentially long-term, adverse neurobehavioral outcomes^34–36^. In this study, we identified an increase in *IL1β* mRNA expression in the fetal side of the placenta in asymptomatic patients, as well as increased expression in maternal blood collected within 14 days of a positive SARS-CoV-2 test. Documentation of IL-1□ protein levels in cord blood and in neonates born to mothers prenatally infected with SARS-CoV-2, as well as newborn testing for SARS-CoV-2 and long-term neurodevelopmental follow-up of such babies, are needed to determine short- and long-term effects of inflammation, infection, or both.

The antiviral response to SARS-CoV-2 includes development of antibodies that recognize the S-RBD as well as neutralize virus ^37^. Detection of anti-SARS-CoV-2 IgG antibodies in maternal and neonatal blood following infection has been reported^38,39^; how symptomatology and pregnancy status, however, affect detection (qualitative) and titers (quantitative) of anti-SARS-CoV-2 IgG and nAb responses has not been previously investigated. Here, we demonstrate that pregnant women infected with SARS-CoV-2 had lower titers of anti-S-RBD IgG compared to non-pregnant women. Although nAb titers were similar between pregnant and non-pregnant women, pregnant women were significantly less likely to have detectable nAb responses. Furthermore, SARS-CoV-2 infected pregnant women who had non-detectable nAb responses had significantly lower anti-S-RBD IgG titers. Reduced antiviral antibody responses in pregnant women infected with SARS-CoV2 were independent of time since infection. The women with low antibody titers did not present with worse symptoms or experience worse disease outcomes, similar to studies in non-pregnant adults^40,41^. It is possible, however, that the reduced antibody titers could increase the potential for reinfection following pregnancy.

Limitations of this study include the small sample size as well as significant differences in age, race, and ethnicity between SARS-CoV-2-infected pregnant and non-pregnant women. These differences are attributable to our reliance on convenience sampling and are a result of differences in participant recruitment, in which sample collection from pregnant women was based on time of delivery, and sample collection from non-pregnant women was based on symptom presentation. While there was a significant difference in age between the cohorts, all women in this study were within reproductive ages^12^. Due to our inability to know precisely when each participant was infected with SARS-CoV-2, we used the number of days between a SARS-CoV-2 PCR test and blood collection as the metric to assess cytokine responses, and additionally used the number of days since symptom onset to evaluate humoral responses over time. These metrics may not accurately represent the time since initial infection, as symptom onset is self-reported and studies have reported PCR positivity for extended periods of time past the initial infection^42,43^.

Our results demonstrate potential differences in the pathogenesis of SARS-CoV-2, including inflammatory and antibody responses to the virus, between pregnant and non-pregnant women. It is well-established that immune responses change dramatically during pregnancy in order to accommodate the developing fetus^44^. Therefore, understanding the impact of SARS-CoV-2 infection during pregnancy on the maternal immune system, and how these changes alter maternal and fetal susceptibility to disease is crucial for the development of vaccines and other therapeutics for COVID-19. Currently, none of the ongoing Phase III trials for promising SARS-CoV-2 vaccine candidates consider pregnant women. In addition to further investigations of short- and long-term consequences of SARS-CoV-2 infection in pregnancy, the safety, immunogenicity, and efficacy of SARS-CoV-2 vaccines in pregnant women must be considered.

## Data Availability

All data are contained in the manuscript.

## Supplemental Materials

1. **eMethods**
2. **eTable 1**
3. **eFigure 1**
4. **References**

## Contributions

IB, SK, AP, JSS, AJS, WCG, and KJ-B conceived of the study and experimental questions, TB, RR, AARA, and YCM collected and provided samples, MJ, TB, DMB, SNW, and RR obtained and organized clinical data, MLS, JL, PC, KL, AP, H-SP, RLU, and AG processed blood samples, MLS, PC, JL, KV, and SO analyzed and graphed data, MLS, MJ, IB, and SK wrote the manuscript, all authors reviewed, edited, and approved the final submission.

## Acknowledgements

The authors thank patients who enrolled and participated in this research and the nurses and staff at the Johns Hopkins Hospitals for assistance with recruitment and sample collection from patients. We thank the National Institute of Infectious Diseases, Japan, for providing VeroE6TMPRSS2 cells and acknowledge the Centers for Disease Control and Prevention, BEI Resources, NIAID, NIH for SARS-related coronavirus 2, isolate USA-WA1/2020, NR-5228. The authors would also like to thank Janna Shapiro for assistance in figure development.

## Data sharing statement

All data are contained in the manuscript.

