## Supplemental Methods, Supplemental Table 1, Supplemental Figure 1 for "Dysregulated immunity in SARS-CoV-2 infected pregnant women"

1. **eMethods**
2. **eTable 1**
3. **eFigure 1**
4. **References**

**1. Methods**

**Study Participants, sample collection, and storage**

**Pregnancy Cohort**

Pregnant women were recruited through Johns Hopkins Hospital outpatient obstetric clinics and the JHH Labor & Delivery unit prior, or after delivery of the patient. We utilized discarded maternal blood, discarded neonatal cord blood, and a small placental sample collected during admission for delivery. Patients were contacted, informed of the study, and consented by phone to decrease face to face exposure due to concern of SARS-CoV-2 spread/infection. Basic demographic information and clinical data, including info on the history of SARS-CoV-2 testing (usually via the POCT nasopharyngeal swab) and symptom expression (asymptomatic or symptomatic), was collected from the patient’s medical record. Blood samples were collected into gold top SST tubes and purple top EDTA tubes. The top SST tubes were inverted several times, before being centrifuged for 10 minutes at 3000 rpm at 22°C. Then, both maternal and cord whole blood and serum samples were aliquoted and stored at -80°C. Placental samples were collected after delivery and were not treated with any preservatives or reagents. Samples were processed using two different methods for both the maternal and fetal sides; placental tissue was either frozen at -80°C immediately or was placed in RNAlater for 48 hours prior to -80°C storage. To obtain tissue that was representative of the placental sample, half thickness samples using a disc tissue punch were taken from two different locations on each side of the placenta. Thus, ultimately, each method of processing placental tissue had two tissue punches from different locations on a given side of the placenta.

**Non-Pregnant Cohort**

A convenience sample of non-hospitalized participants were recruited and provided informed consent by phone between April 21 and August 13, 2020 after receiving a positive SARS-CoV-2 RT-PCR test from an outpatient or emergency department facility within the Johns Hopkins Health Sytem^1^. One participant requested participation in the study via the Johns Hopkins HOPE (Hopkins Opportunities for Participant Engagement) COVID-19 registry. Samples from adult women of reproductive age, 18-49 years ^2^, with positive RT-PCR results for SARS-CoV-2 were included in this study. Basic demographic information and clinical data, including that regarding the history of SARS-CoV-2 testing, was collected from the patient and the patient’s medical record. Participants in this study attended a research clinic visit on average 42.2 days after COVID-19 symptom onset (range 29-92 days), at which blood was drawn. Approximately 25 ml of whole blood was collected in ACD tubes. Peripheral blood mononuclear cells were separated, and the remaining plasma was stored in 1 ml aliquots at -80^o^C. Plasma was defrosted and then heat inactivated at 56^o^C for 30 minutes prior to serologic assays. The study was approved by the Johns Hopkins School of Medicine Institutional Review Board.

**Gene Expression Analysis**

Total RNA was extracted from placental tissue samples using the RNeasy Plus Mini Kit (Qiagen) or from whole blood using NucleoSpin RNA Blood Kit (Macherey-Nagel). Complementary (c) DNA synthesis in a 40‐μL reaction was performed using Bio‐Rad iScript™ cDNA Synthesis Kit (Bio‐Rad). TaqMan® (Thermo Fisher Scientific) mRNA assays were run for analysis. The primers used were IL-1β (Hs.PT.58.1518186; Integrated DNA Technologies) and IL-6 (Hs.PT.58.40226675; Integrated DNA Technologies). mRNA expression was calculated relative to housekeeping genes: 18S (Applied Biosystems) and Actin (Hs. PT.39a.22214847; Integrated DNA Technologies).

**Indirect enzyme-linked immunosorbent assays (ELISAs)**

The protocol was adapted from a published protocol from Dr. Florian Krammer’s laboratory^3^, as described in Klein et al., 2020^4^. Briefly, ninety-six-well plates (Immulon 4HBX, Thermo Fisher Scientific) were coated with either full-length S protein or S-RBD at 4^o^C overnight. Coating buffer was removed, and plates were washed and then blocked for 1 hour at room temperature. All plasma samples were heat inactivated at 56^o^C on a heating block for 1 hour before use. Negative control samples were prepared at 1:10 dilutions and plated at a final concentration of 1:100. A mAb against the SARS– CoV-2 S protein was used as a positive control (1:5000; catalog 40150-D001, Sino Biological). For serial dilutions of plasma on either S- or S-RBD–coated plates, plasma samples were prepared in 3-fold serial dilutions starting at 1:20. Blocking solution was removed, and 10 μL diluted plasma was added in duplicate to the plates and incubated at room temperature for 2 hours. Plates were washed 3 times with PBST wash buffer, and 50 μL secondary antibody was added to the plates and incubated at room temperature for 1 hour (Fc-specific total IgG HRP 1:5000 dilution, catalog A18823, Invitrogen, Thermo Fisher Scientific). Plates were washed and all residual liquid removed before addition of 100 μL SIGMAFAST OPD (o phenylenediamine dihydrochloride) solution (MilliporeSigma) to each well, followed by incubation in darkness at room temperature for 10 minutes. To stop the reaction, 50 μL 3M HCl (Thermo Fisher Scientific) was added to each well. The OD of each plate was read at 490 nm (OD490) on a SpectraMax i3 ELISA Plate Reader (BioTek Instruments). The positive cutoff value for each plate was calculated by summing the average of the negative values and 3 times the SD of the negatives. All values at or above the cutoff value were considered positive.

**Microneutralization assay**

The plasma neutralizing antibody (nAb) protocol was adapted from Dr. Andrew Pekosz’ laboratory^5^, as described in Klein et al., 2020^4^. Briefly, infectious virus (SARS-CoV-2/USA-WA1/2020) was added to two-fold diluted plasma at a final concentration of 1 × 104 TCID50/mL (100 TCID50 per 100 μL). Samples were added to VeroE6-TMPRSS2 cells in sextuplet for 6 hours at 37˚C. The inocula were removed, fresh IM was added, and the plates were incubated at 37˚C for 2 days. Cells were fixed by the addition of 150 μL of 4% formaldehyde per

well, incubated for at least 4 hours at room temperature, and then stained with Napthol Blue Black (MilliporeSigma). The nAb titer was calculated as the highest serum dilution that eliminated the cytopathic effect in 50% of the wells.

**Western blot**

Western blotting was used to measure the protein expression of FcRn in placenta. To prepare tissue lysate, tissue was homogenized on ice in RIPA lysis buffer (Sigma‐Aldrich) with proteinase inhibitor (Sigma‐Aldrich) and phosphatase inhibitor cocktail 2 (Sigma‐Aldrich). The homogenized specimens were then placed on ice for 15 min and centrifuged at 14,000 rpm for 20 min at 4°C. The resulting supernatants were collected for further experiments. Total protein was separated by sodium dodecyl sulfate‐polyacrylamide gel electrophoresis (SDS‐PAGE, Bio‐Rad) using 4%–15% gels (Bio‐Rad), and then, transferred onto nitrocellulose membranes (Bio‐Rad) using semidry transfer device (Trans‐Blot® Turbo™, Bio‐Rad). Membranes were blocked with 5% of bovine serum albumin (BSA, Sigma‐Aldrich) in Tris‐buffered saline (Corning) plus 0.1% of Tween‐20 (Sigma‐ Aldrich) (TBST) for 15 min at room temperature, and incubated with primary antibodies in 5% of BSA at 4°C overnight, then, washed using TBST. FcRn antibody (1:1000, Santa Cruz) and GAPDH (control marker, 1:1000, Abcam) were used for primary antibodies. ECL (GE Healthcare) was used for detection using the ImageQuant LAS 500 (GE Healthcare), and densitometric analysis was performed using ImageJ (National Institutes of Health; http://rsb.info.nih.gov/ij/).

**Statistical Analysis**

Descriptive statistics stratified by pregnancy state (SARS-CoV-2 positive pregnant, SARS-CoV-2 positive non-pregnant) and symptomatology (symptomatic or asymptomatic SARS-CoV-2 positive pregnant) are presented as medians and IQRs. Comparisons of demographic characteristics were tested via exact Wilcoxon two-sample test, Pearson’s chi-squared test, or Fisher’s exact test, where appropriate dependent on variable structure as continuous, binary, or categorical and sample size within individual cells. Prior to conducting any inferential statistics, AUC values for anti-S IgG and anti-S-RBD IgG titers were computed by plotting normalized OD values against sample dilution for ELISAs. The AUC for microneutralization assays used the exact number of wells protected from infection at each plasma dilution. For each assay, samples with titers below the limit of detection were assigned an AUC value of half of the lowest measured AUC value. Due to the non-normal distribution of cytokine and antibody data, comparisons between symptomatology among pregnant women, as well as, comparison between SARS-CoV-2 positive pregnant and SARS-CoV-2 positive non-pregnant women were examined via exact Wilcoxon two-sample tests. Comparison across three groups (SARS-CoV-2 symptomatic positive pregnant, SARS-CoV-2 asymptomatic positive pregnant, SARS-CoV-2 negative pregnant) were tested via a Kruskal-Wallis test exact, followed by pairwise multiple comparisons. Correlations between antibody isotypes and assays with days since the initial SARS-CoV-2 positive test or days since symptom onset were assessed using the Spearman correlation coefficient. The data were then log transformed for visualization. Finally, a generalized linear model was used to determine if the association between days since the initial SARS-CoV-2 positive test or days since symptom onset and antibody responses differed by pregnancy status (pregnant, non-pregnant). All analyses were two-tailed tests with a significance threshold of *p* < .05.

**2. eTable 1. Comparison of clinical and pregnancy outcome data between SARS-CoV-2 positive (+) and negative (-) pregnant women.**

| **Pregnant Female Cohort** | **All** | **SARS-CoV-2 (+)** | **SARS-CoV-2 (-)** | **P value *** |
| --- | --- | --- | --- | --- |
| **N (%)** | 33 | 22 (66.67) | 11 (33.33) |  |
| **Median BMI (pre-pregnancy)** | 24.99 | 26.3 | 23.67 | **0.2713** |
| **Median BMI (at delivery)** | 29.48 | 29.8 | 28.08 | **0.144** |
| **Median gestational age at delivery** | 39.1 | 38.85 | 39.5 | **0.0725** |
| **Median neonate late-onset sepsis (LOS)** | 2 | 2 | 2 | **0.4451** |
| **Chorioamnionitis** | 2 | 2 | 2 | **0.5** |
| **Membrane rupture >18 hours prior to delivery** | 4 | 4 | 0 | **0.132** |
| **Preeclampsia-n(%)** |  |  |  |  |
| Yes | 2(6.06) | 2(9.09) | 0(0) | **0.5417** |
| No | 31(93.94) | 20(90.91) | 11(100) |  |
| **Gestational Diabetes-n(%)** |  |  |  |  |
| Yes | 2(6.06) | 2(9.09) | 0(0) | **0.5417** |
| No | 31(93.94) | 20(90.91) | 11(100) |  |
| **Delivery Type-n(%)** |  |  |  |  |
| Cesarean | 12(36.36) | 7(31.82) | 5(45.45) | **0.4713** |
| Vaginal | 21(63.64) | 15(68.18) | 6(54.55) |  |
| **Size of neonate-n(%)** |  |  |  |  |
| AGA | 26(78.79) | 17(77.27) | 9(81.82) | **1** |
| LGA | 5(15.15) | 3(13.64) | 2(18.18) |  |
| SGA | 2(6.06) | 2(9.09) | 0(0) |  |
| **Sex of neonate-n(%)** |  |  |  |  |
| Female | 18(54.55) | 13(59.09) | 5(45.45) | **0.4583** |
| Male | 15(45.45) | 9(40.91) | 6(54.55) |  |
| **NICU stay-n(%)** |  |  |  |  |
| Yes | 4(12.12) | 4(18.18) | 0(0) | **0.2755** |
| No | 29(87.88) | 18(81.82) | 11(100) |  |
| **Neonate readmission-n(%)** |  |  |  |  |
| Yes | 1(3.03) | 0(0) | 1(9.09) | **0.3333** |
| No | 32(96.97) | 0(0) | 10(90.91) |  |

**3. eFigure 1.** ***IL6* expression in maternal and fetal samples.** Maternal and fetal blood and placentas were used to detect *IL6* gene expression relative to the housekeeping genes (HKG), *18S* and *ACTB*. (**A-D**) Maternal blood, cord blood, and maternal and fetal side placental *IL6* expression between SARS-CoV-2 positive (P(+)) and negative (P(-)) samples in the pregnant cohort. (**E-H**) Maternal blood, cord blood, and maternal and fetal side placental *IL6* expression in pregnant women who were asymptomatic (P-A), symptomatic (P-S), or SARS-CoV-2 negative (P-N). (**I**) Maternal blood *IL6* expression analyzed as a function of symptom expression and days between SARS-CoV-2 PCR positive test and blood sample collection; dashed line located at Day 14. Maternal blood n= 27; cord blood=29; maternal side placenta n=11; fetal side placenta n=26.

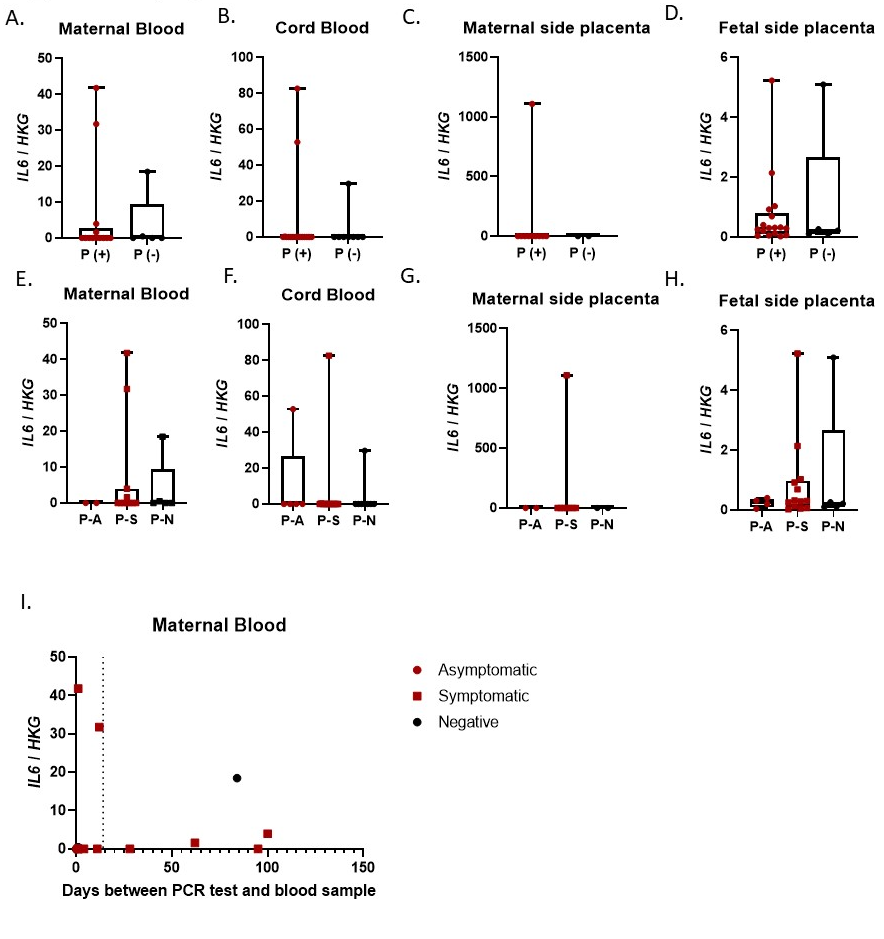
